# Diverging functional connectivity timescales: Capturing distinct aspects of cognitive performance in early psychosis

**DOI:** 10.1101/2024.05.07.24306932

**Authors:** Fabian Hirsch, Ângelo Bumanglag, Yifei Zhang, Afra Wohlschlaeger

**Affiliations:** Department of Diagnostic and Interventional Neuroradiology, Klinikum R.d.Isar, Technical University Munich, Ismaninger Str. 22, Munich 81675, Germany

**Keywords:** {MRI, resting-state, time-varying connectivity, early psychosis, cognition

## Abstract

**Background:** Psychosis spectrum disorders (PSDs) are marked by cognitive impairments, the neurobiological correlates of which remain poorly understood. Here, we investigate the entropy of time-varying functional connectivity (TVFC) patterns from resting-state fMRI (rfMRI) as potential biomarker for cognitive performance in PSDs. By combining our results with multimodal reference data, we hope to generate new insights into the mechanisms underlying cognitive dysfunction in PSDs. We hypothesized that low-entropy TVFC patterns (LEN) would be more behaviorally informative than high-entropy TVFC patterns (HEN), especially for tasks that require extensive integration across diverse cognitive subdomains.

**Methods:** rfMRI and behavioral data from 97 patients in the early phases of psychosis and 53 controls were analyzed. Positron-Emission Tomography (PET) and magnetoencephalography (MEG) data were taken from a public repository (Hansen et al., 2022). Multivariate analyses were conducted to examine relationships between TVFC patterns at multiple spatial scales and cognitive performance in patients.

**Results:** Compared to HEN, LEN explained significantly more cognitive variance on average in PSD patients, driven by superior encoding of information on psychometrically more integrated tasks. HEN better captured information in specific subdomains of executive functioning. Nodal HEN-LEN transitions were spatially aligned with neurobiological gradients reflecting monoaminergic transporter densities and MEG beta power. Exploratory analyses revealed a close statistical relationship between LEN and positive PSD symptoms.

**Conclusion:** Our entropy-based analysis of TVFC patterns dissociates distinct aspects of cognition in PSDs. By linking topographies of neurotransmission and oscillatory dynamics with cognitive performance, it enhances our understanding of the mechanisms underlying cognitive deficits in PSDs.

**CRediT Authorship Contribution Statement:** **Fabian Hirsch:** Conceptualization, Methodology, Software, Formal analysis, Writing - Original Draft, Writing - Review & Editing, Visualization; **Ângelo Bumanglag:** Methodology, Software, Formal analysis, Writing - Review & Editing; **Yifei Zhang:** Methodology, Software, Formal analysis, Writing - Review & Editing; **Afra Wohlschlaeger:** Methodology, Writing - Review & Editing, Supervision, Project administration

## 1 Introduction

Psychosis spectrum disorders (PSDs) are marked by positive and negative symptoms, as well as cognitive impairment (McTeague et al., 2017; R. M. Murray et al., 2004; Pearlson, Clementz, Sweeney, Keshavan, & Tamminga, 2016; Sharma et al., 2017). PSDs encompass traditionally distinct diagnostic categories like schizophrenia (SCZ) and bipolar disorder (BP) (Barch, 2017; Yamada, Matsumoto, Iijima, & Sumiyoshi, 2020), with positive symptoms like hallucinations and delusions being the predominant feature across these categories (van Os & Kapur, 2009). Positive symptoms have traditionally received a lot of attention in research on PSDs (Feinberg, 1978; Kapur, 2003; Sterzer et al., 2018), while cognitive dysfunction is less frequently discussed (Harvey et al., 2022). However, cognitive impairment is predictive of developing psychosis in high-risk individuals (Carrion et al., 2016; Seidman et al., 2016) and is a core feature across different manifestations of PSD (Bora & Pantelis, 2015). Consequently, mapping the neurophysiological correlates of cognitive performance in PSDs is an important subject of investigation, especially since the therapeutic effects of antipsychotic medications on cognitive deficits are merely moderate (Keefe et al., 2007), or even entirely absent for certain substances (Baldez et al., 2021). This mapping can be performed with functional magnetic resonance imaging (fMRI), where functional connectivity (FC) changes across large-scale brain networks (NWs) in PSD patients have been reported (Anticevic et al., 2014; Cheng et al., 2015; Kambeitz et al., 2016; Ramsay, 2019; N. D. Woodward & Heckers, 2016). These changes are said to reflect dysfunctional integration of information between different brain systems with distinct roles in the processing hierarchy (Anticevic & Halassa, 2023; Friston, Brown, Siemerkus, & Stephan, 2016), ultimately giving rise to the diverse set of PSD symptoms.

Besides FC, which is usually measured by the correlation between two blood-oxygen-level-dependent (BOLD) signals during rest, intrinsic properties of BOLD timeseries also reflect integrative processes (Garrett, Epp, Perry, & Lindenberger, 2018; Ito, Hearne, & Cole, 2020). The related concept of intrinsic neural timescales (INT) suggests that more self-similarity (longer INT) in a local BOLD signal reflects a longer temporal window for the integration of information within that brain region (Hasson, Chen, & Honey, 2015; Stephens, Honey, & Hasson, 2013). Fittingly, INTs have been shown to be significantly shortened in PSD patients compared to healthy controls (Uscatescu et al., 2023; Uscatescu et al., 2021; Wengler, Goldberg, Chahine, & Horga, 2020), and follow a spatial gradient from primary-sensory to higher order regions (J. D. Murray et al., 2014; Raut, Snyder, & Raichle, 2020). Relevant to the present investigation, the concept of INT can be extended to the level of connections (edges), by focusing on time-varying aspects of FC (TVFC). Although TVFC is still a controversial topic (Liegeois, Laumann, Snyder, Zhou, & Yeo, 2017; Lurie et al., 2020), studying it has proven to be informative regarding interindividual differences (Liegeois et al., 2019; Vidaurre, Llera, Smith, & Woolrich, 2021) and disease states (Jia, Gu, & Luo, 2017; Kaiser et al., 2016; Sakoglu et al., 2010). Consequently, a small number of resting-state fMRI studies have used sample entropy (SampEn) (Richman & Moorman, 2000) to quantify the self-similarity of edge fluctuations (edge-SampEn [ESE]) derived from sliding-window analysis (Hirsch & Wohlschlaeger, 2022; Jia & Gu, 2019b; Jia et al., 2017). SampEn is one way of assessing INT, with higher values corresponding to shorter INT (Omidvarnia, Mesbah, Pedersen, & Jackson, 2018; Sokunbi et al., 2014), and it is also significantly associated with mental abilities and cognitive load in healthy subjects (S. S. Menon & Krishnamurthy, 2019; Nezafati, Temmar, & Keilholz, 2020; Omidvarnia et al., 2022; Omidvarnia et al., 2021).

Evidence further indicates that ESE can provide complementary information to BOLD-derived SampEn (S. S. Menon & Krishnamurthy, 2019), suggesting potential use as a novel biomarker for neuropsychiatric conditions and their related symptoms. In support of this hypothesis, Jia and Gu (2019a) reported that ESE was significantly higher in SCZ patients at multiple spatial scales compared to healthy controls. However, statistical relationships between ESE and cognitive task-performance have not been explored in PSDs. Open questions also pertain to the possibly differential contributions of high and low ESE configurations to performance in patients: High ESE connections were most predictive of fluid intelligence in healthy subjects (S. S. Menon & Krishnamurthy, 2019). However, brain regions belonging to cortical NWs associated with visuospatial and language functions display the lowest ESE in the brain (Hirsch & Wohlschlaeger, 2022), and connectivity patterns of these NWs have been repeatedly associated with cognitive ability (Hearne, Mattingley, & Cocchi, 2016; Song et al., 2008; van den Heuvel, Stam, Kahn, & Hulshoff Pol, 2009). To address these issues, we analyze resting-state fMRI and behavioral data from a clinical population (*n* = 97) of young adults that is within 3 years of onset of psychotic symptoms, as well as from healthy controls (*n* = 53). We contrast high and low ESE network configurations, in terms of their ability to explain behavioral variance across cognitive tasks in patients. Given the evidence cited above, we hypothesize that their respective explanatory power would significantly depend on the specific cognitive task in question: Low ESE configurations should be more informative in tasks that need higher degrees of information integration. Conversely, high ESE configurations might better capture behavioral variance in tasks that depend more on ‘just’ the precise encoding of low-level stimulus features. Overall, we hope to generate new perspectives regarding the topography of neurophysiological correlates of cognitive performance in PSDs, through examining the timescales of TVFC with ESE. By combining our fMRI results with public data of neurotransmitter systems and brain oscillations (Hansen et al., 2022), we aim to gain more insight into the biological mechanisms underlying ESE configurations and their relationship with cognitive aspects of PSDs. This multimodal mapping of brain-behavior associations might help to generate new potential targets for therapeutic interventions in the cognitive domain, particularly since existing treatment options are only moderately effective (Vita et al., 2021).

## 2 Results

Imaging and behavioral data were taken from the Human Connectome Project for Early Psychosis (HCP-EP) open-source dataset (Materials and Methods). Imaging data consisted of one resting-state session (*∼*6min) per subject (patients: *n* = 97; controls: *n* = 53). Behavioral data consisted of scores from the seven measures in the NIH-TB Cognition Battery (Weintraub et al., 2013), that capture individual variation across a range of cognitive subdomains. To map brain-behavior relationships in patients, we used multi- and univariate versions of a variance component model (Ge et al., 2016; Sabuncu et al., 2016), that has been recently employed to study TVFC - behavior associations in healthy subjects (Liegeois et al., 2019). After preprocessing, the functional data were parcellated and sliding-window analysis (SWA) was conducted on the resulting BOLD timeseries. One SampEn value was then computed for each correlational timeseries, resulting in a vector with 6670 elements for each subject. In accordance with previous work, we then constructed high-entropy (HEN) and low-entropy (LEN) network-templates, by selecting edges with the highest and lowest mean ESE values across healthy subjects (Hirsch & Wohlschlaeger, 2022). This was done for a range of different thresholds, and for every threshold we extracted the corresponding ESE values from the patients, which were then correlated across patients to derive the similarity matrices to be put into the model (Ge et al., 2016). We then ran the behavioral model for each similarity matrix corresponding to a given threshold, and in the end selected the threshold that performed best for HEN and LEN (respectively) for the final analysis (Section 4.3). The two resulting (97 x 97) similarity matrices *RHEN* and *RLEN* (representing shared variance in ESE across patients) were then used as separate inputs for the variance component model to predict variance across and within cognitive domains. All ensuing behavioral analyses are based on comparing the outcomes from running the model separately for *RHEN* and *RLEN* (Materials and Methods).

Moreover, we performed basic topological analyses at the node-level, based on binarized versions of the (group-level) HEN and LEN templates, derived from the controls. This was done to replicate our previous finding that ESE is topographically organized along a subcortical (SC) to cortical axis in healthy subjects (Hirsch & Wohlschlaeger, 2022). Finally, to gain more insight into the neurobiological mechanisms underlying HEN and LEN configurations at the cortical level, we analyze their spatial correspondence with neurotransmitter maps derived from positron emission tomography (PET) and the topography of brain rhythms from magnetoencephalography (MEG). We use high-quality open-source data that combines the results from different studies (Hansen et al., 2022). We apply rigorous control for statistical dependencies between spatially adjacent brain regions through the employment of null-models matching the spatial autocorrelation of the empirical maps (Burt, Helmer, Shinn, Anticevic, & Murray, 2020).

### 2.1 LEN encodes more information across cognitive domains in patients

On average, similarity in the HEN explained significantly less behavioral variance (26%; *SE* = 16%), compared to the LEN (36%; *SE=* 20%), bias-corrected bootstrap confidence-interval (BS-CI) of the difference [-26%, -4%], Bonferroni corrected (Figures 1-2). There was reasonable evidence for the average explanatory variance to be significantly different from zero for both LEN (*p*-Wald = 0.0387,*p*-Perm = 0.034), and HEN (*p* -Wald = 0.0443, *p* -Perm = 0.045). Running the model after shuffling the original edges (independently for each patient) or selecting random edges (consistently across patients) resulted in higher p-values for both HEN (shuffled-edges: *p* -Wald = 0.4477, *p* -Perm = 0.4270; random edges: *p* -Wald = 0.1602, *p* -Perm = 0.1560) and LEN (shuffled-edges: *p* -Wald = 0.4858, *p* -Perm = 0.4610; random-edges: *p* -Wald = 0.1665, *p* -Perm = 0.1650).

**Figure 1.**
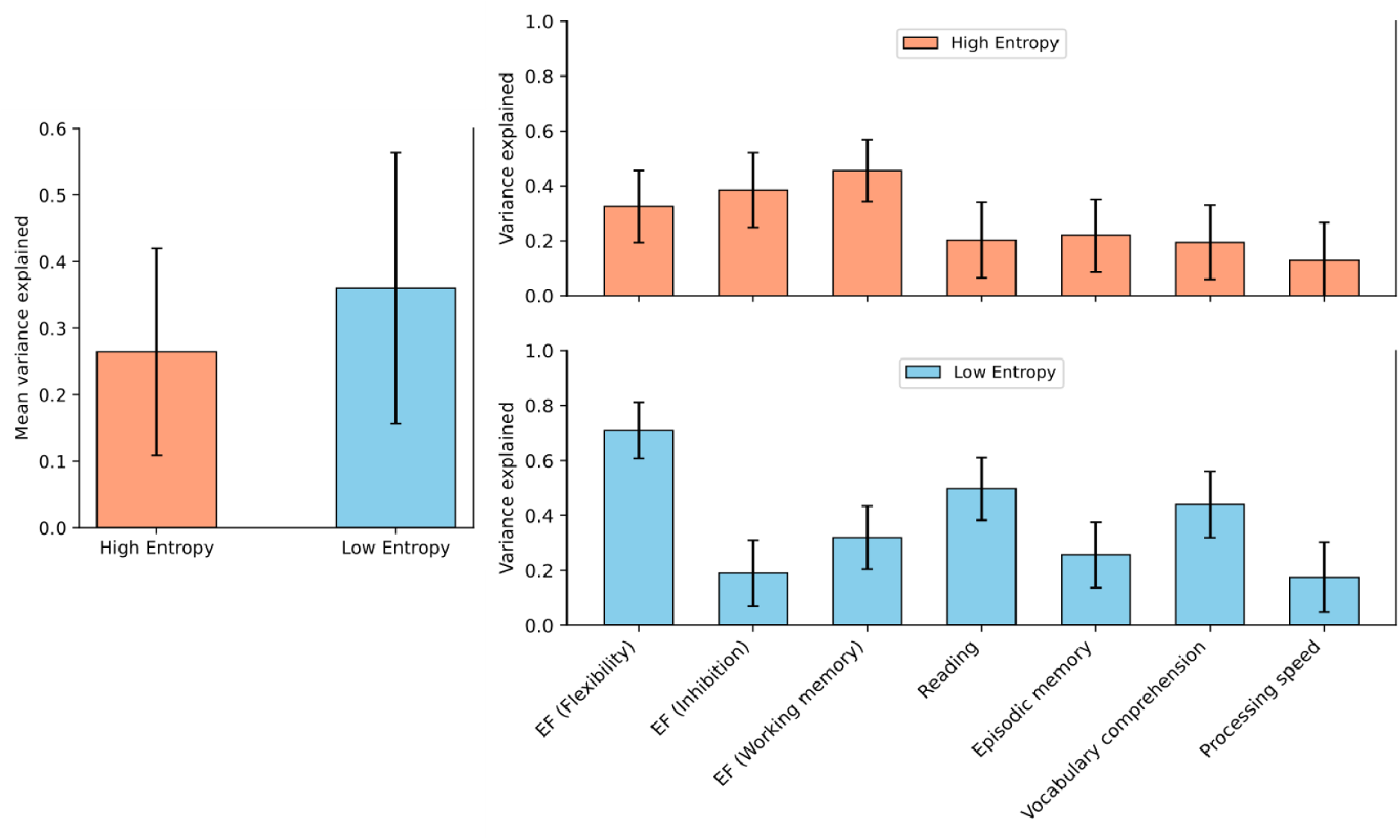
*<u>Left:</u>* Average variance explained across all cognitive measures is significantly lower for the High-entropy (red), compared to the Low-entropy (blue), in patients. Error-bars represent parametric SEs. *<u>Right:</u>* Explained variance in patients, stratified by cognitive measure for High-entropy (top-row) and Low-entropy (bottom-row). Error bars represent SEs derived from a bootstrapping procedure. SE= Standard error, EF = Executive functions.

### 2.2 Explanatory power is domain specific

On the level of single measures, we observed significant interactions in subdomains of Executive Function (EF): HEN (39%; SE= 14%) explained more variance in Inhibition (INH), compared to LEN (19%; SE= 12%), BS-CI [16%, 66%]. HEN (46%; SE = 11%) also explained significantly more variance in Working-memory (WM), compared to LEN (32%; SE = 12%), BS-CI [9%, 49%]. Conversely, HEN (33%; SE= 13%) explained significantly less variance in Flexibility, compared to LEN (71%; SE = 10%), BS-CI [-92%, -48%] (Figure 2). Additionally, HEN was significantly less informative in Reading (20%; SE= 14%), compared to LEN (50%; SE= 11%), BS-CI [-76%, -29%]. Finally, HEN was also significantly less informative in Vocabulary comprehension (20%; SE= 14%), compared to LEN (44%; SE= 12%), BS-CI [-65%, -22%]. All BS-Cis were (Bonferroni) adjusted for multiple comparisons, and the results were insensitive to the choice of resampling method (Section 4.6).

**Figure 2.**
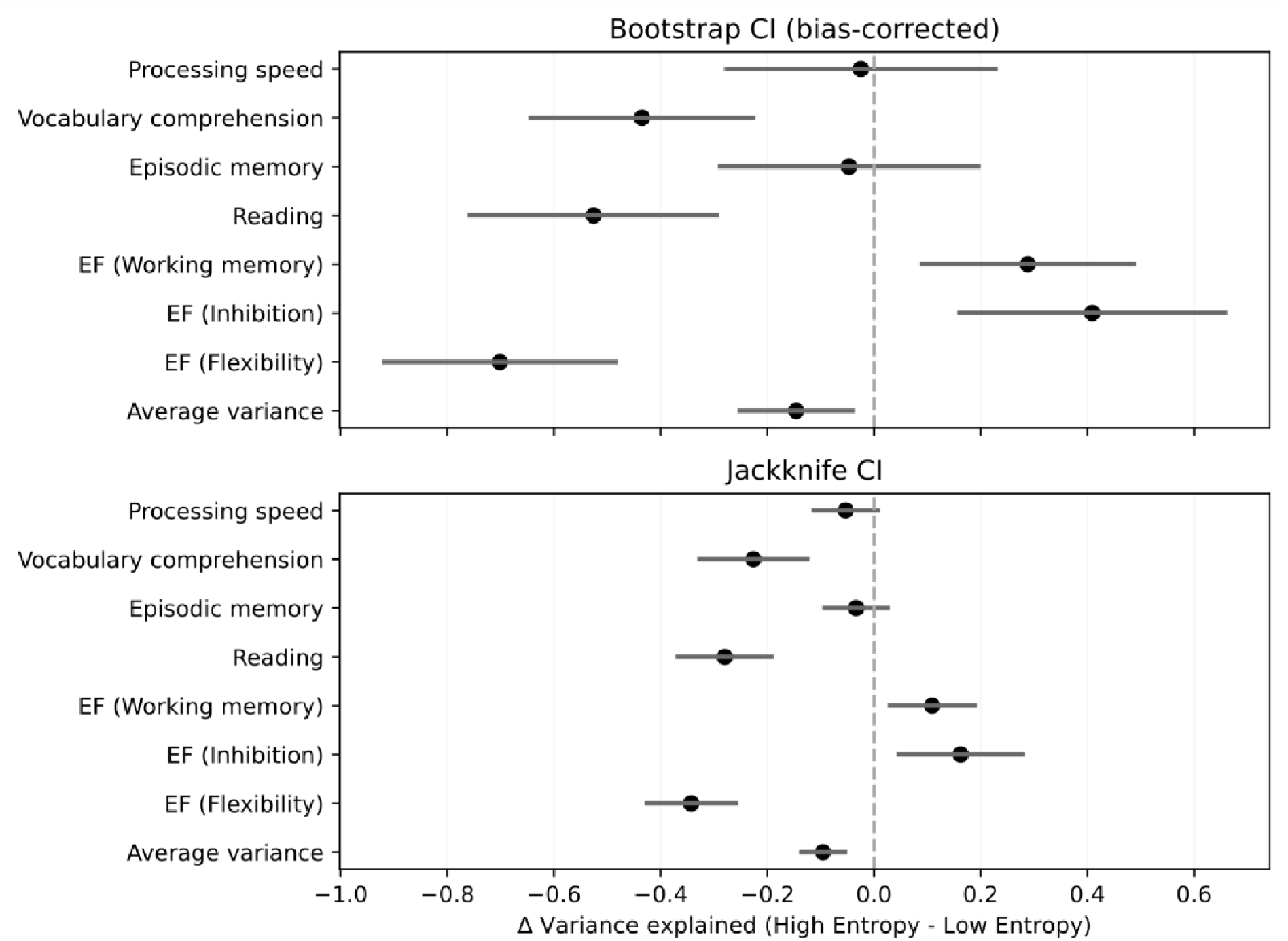
*<u>Top:</u>* Bias-corrected bootstrap Cls for the patients’ difference in variance explained (High-entropy - Low entropy), stratified by cognitive measure. Dashed line denotes zero-difference. *<u>Bottom:</u>* Confirmatory jackknife Cls for the patients’ difference in variance explained (High-entropy - Low-entropy), stratified by cognitive measure. Dashed line denotes zero-difference.Cl = Confidence interval, EF = Executive functions.

### 2.3 Explanatory power is NW specific

To evaluate behavioral variance explained for HEN and LEN at the level of NWs, a univariate version of the multivariate variance component model was used, which resulted in an edgewise estimate quantifying the average amount of variance explained across all dependent variables (Material and Methods). We averaged edges-values within- and between the boundaries of seven established cortical resting-state NWs (Yeo et al., 2011), plus a SC NW. The cortical NWs pertained to the Visual NW (VIS), Somatomotor NW (SM), Dorsal Attention NW (DAT), Salience/Ventral Attention NW (SAL), Limbic NW (LIM), Cognitive Control NW (CC), as well as Default-mode NW (DMN). To determine significance, we compared these empirical values against a series of values derived from 10000 iterations of different null-models (Materials and Methods). We found that for the HEN, the average variance explained within SC was significantly higher than what would be expected based on a series of size- and density-matched random NWs (*p* -Perm = 0.0014) (Figure 3; top left). The same was true for SC interactions with VIS (*p* -Perm = 0.0014), LIM (*p* -Perm = 0.0014), CC (*p* -Perm = 0.0257), and DMN (*p* -Perm = 0.0014) (Figure 3; top left). When compared to degree- and strength-matched random NWs, only SC interactions with CC (*p* -Perm = 0.0072) remained significant (Figure 3; bottom left). For the LEN, the average variance explained within DMN was significantly higher than what would be expected based on series of size and density-matched random NWs (*p*-Perm = 0.0261), as well as DMN interactions with DAT (*p* -Perm = 0.0456) (Figure 3; top right). The same was true for SM interactions with SAL (*p* -Perm = 0.0014), and DAT interactions with CC (*p* -Perm = 0.0252) (Figure 3; top right). When compared to degree- and strength-matched random NWs, no NW (or between-NW interaction) in the LEN explained significantly more behavioral variance across behavioral measures than expected (on average) (Figure 3; bottom right). All reported p-values were controlled with FDR (Benjamini & Hochberg, 1995)

**Figure 3.**
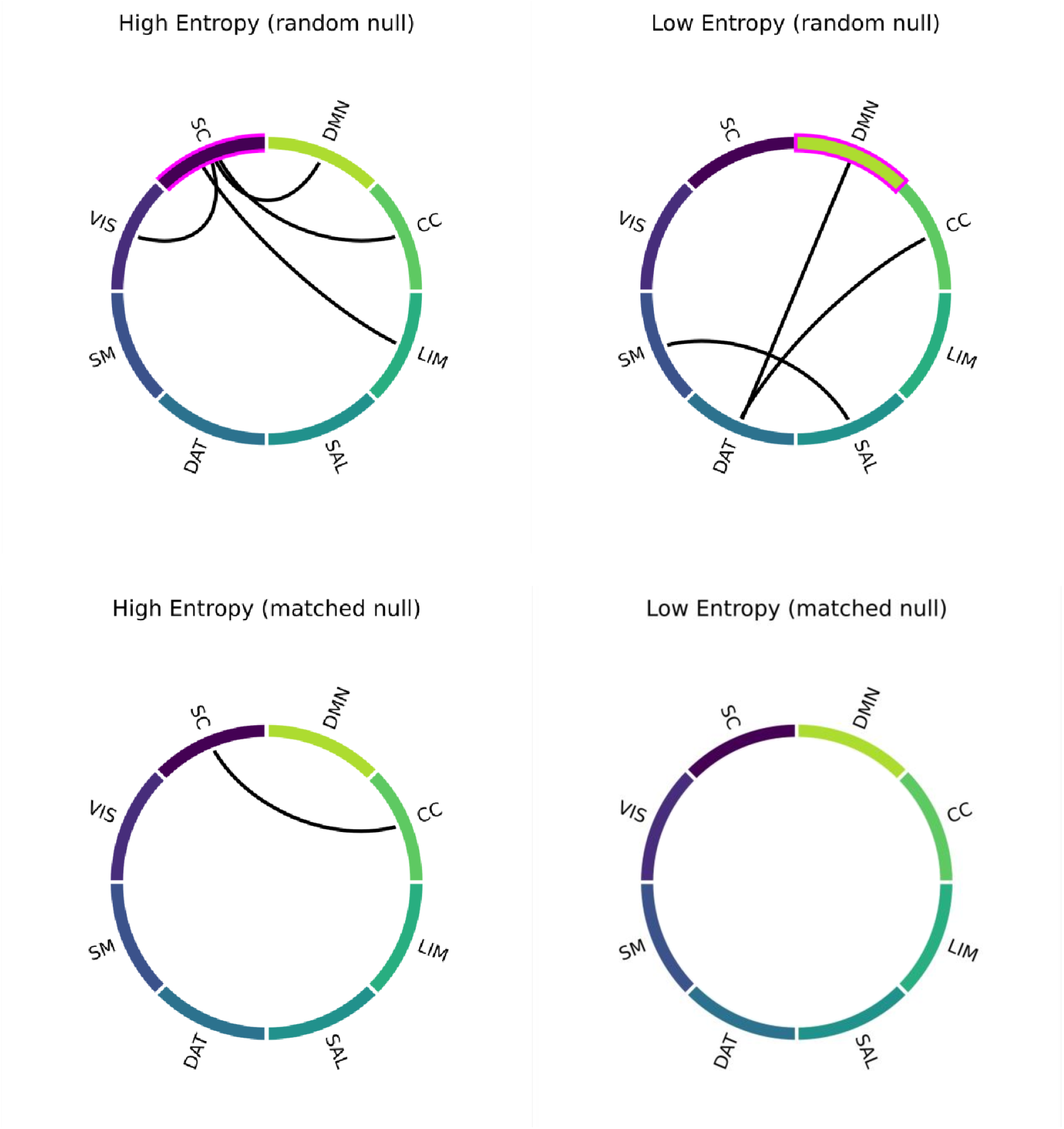
*<u>Top-row:</u>* NW-interactions explaining significant behavioral variance across all cognitive measures in patients, for High-entropy (left) and Low-entropy (right). Significance was determined by randomly shuffling edges (random-null; 10000 permutations) before averaging within/between NWs. Significant within-NW explanatory variance is denoted by magenta colors. *<u>Bottom-row:</u>* NW-interactions explaining significant behavioral variance across all cognitive measures in patients, for High-entropy (left) and Low-entropy (right). Significance was determined by shuffling edges while matching the initial degree- and strength distributions (matched-null; 10000 permutations) before averaging within/between NWs. CC = Cognitive control NW, DAT= Dorsal attention NW, DMN = Default-mode NW, LIM= Limbic NW, SAL= Salience NW, SC= Subcortical NW, SM= Sensorimotor NW, VIS= Visual NW, NW= Network.

### 2.4 Spatial layout of ESE recapitulates SC-cortical axis in controls

We assessed the relative importance of single regions to HEN and LEN configurations by computing the (binary) degree centrality (DC) for each node in the respective templates (Section 2). Each node’s DC value was normalized by the mean DC value from a series of size- and density-matched random NWs. For the HEN, highest DC values were localized in SC, with left hemispheric nodes in the posterior thalamus, amygdala, and hippocampus at the top (Figures 4-5). Cortical nodes with the highest DC values were found in LIM regions of the temporal lobes (bilaterally), as well as in areas belonging to VIS (Figure 6). For the LEN, highest DC values belonged to pre- and postcentral SM and DAT regions (bilaterally), as well as to bilateral prefrontal- and cingulum areas of the CC (Figure 6). Overall, the DC values were spatially organized in strong correspondence with our previous results in large sample of young and healthy subjects (Hirsch & Wohlschlaeger, 2022).

**Figure 4.**
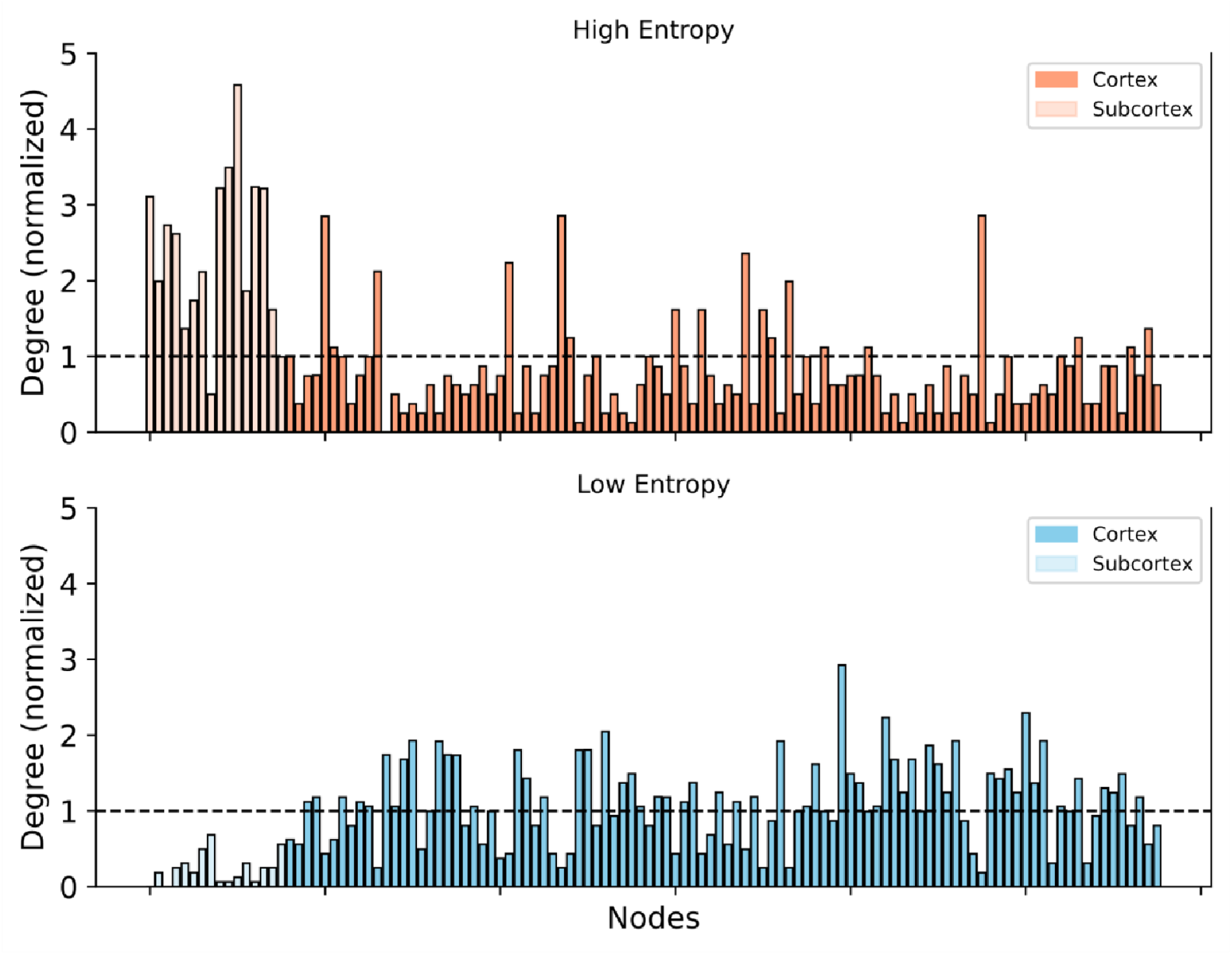
*<u>Top-row:</u>* Normalized degree-centrality of all nodes for High-entropy (red), stratified by localization (Cortex = dark red; Subcortex = light red). Normalization was done via randomly shuffling edges (10000 permutations), dashed line denotes significance. *<u>Bottom-row:</u>* Normalized degree-centrality of all nodes for Low-entropy (blue), stratified by localization (Cortex= dark blue; Subcortex = light blue). Normalization was done via randomly shuffling edges (10000 permutations), dashed line denotes significance.

**Figure 5.**
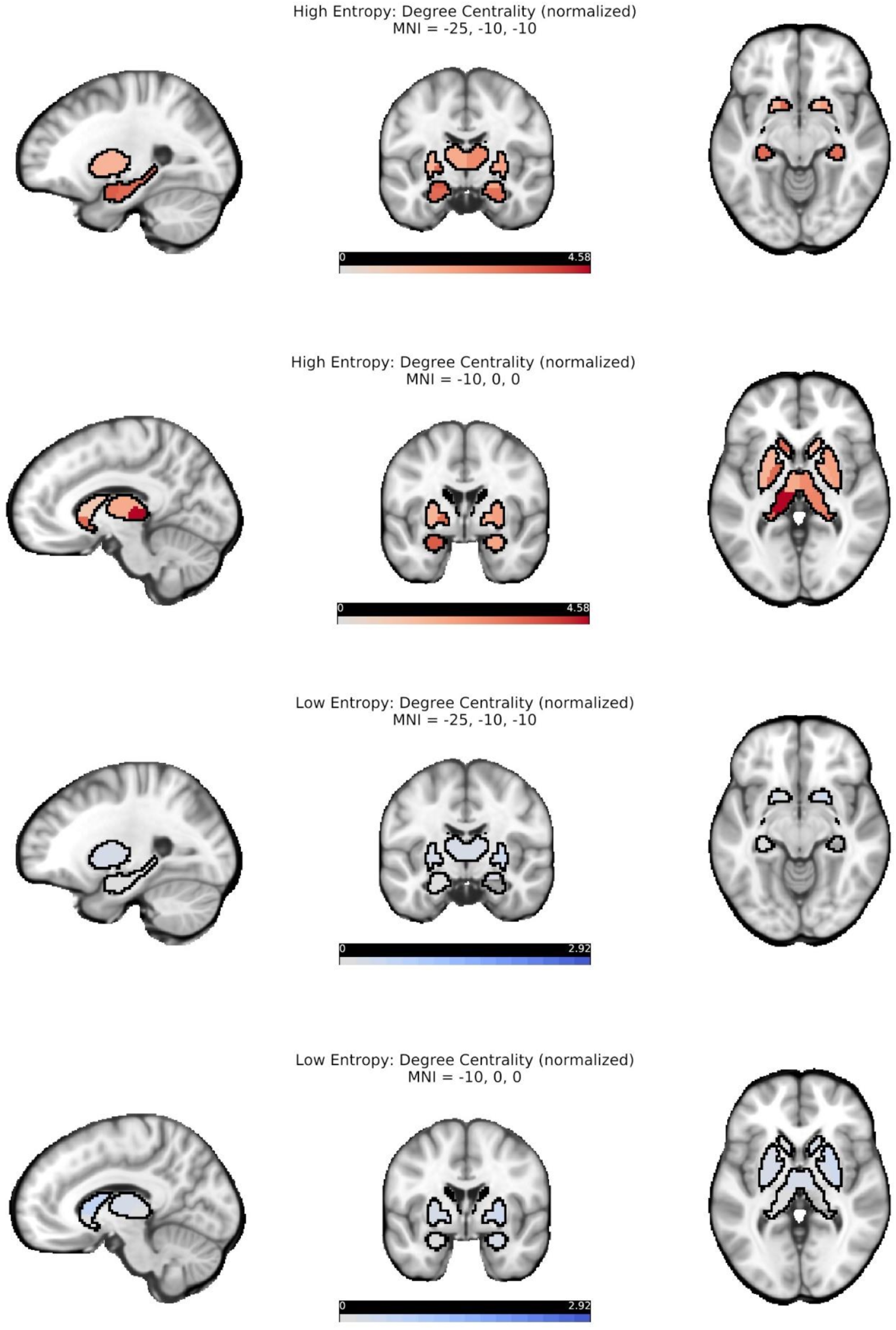
Normalized degree-centrality of subcortical nodes for High-entropy (red) and Low-entropy (blue), depicted on representative slices of a structural image in MNI space. Normalization was done via randomly shuffling edges (10000 permutations), darker colors denote higher degree-centrality. Data are the same as in Figure 4.

**Figure 6.**
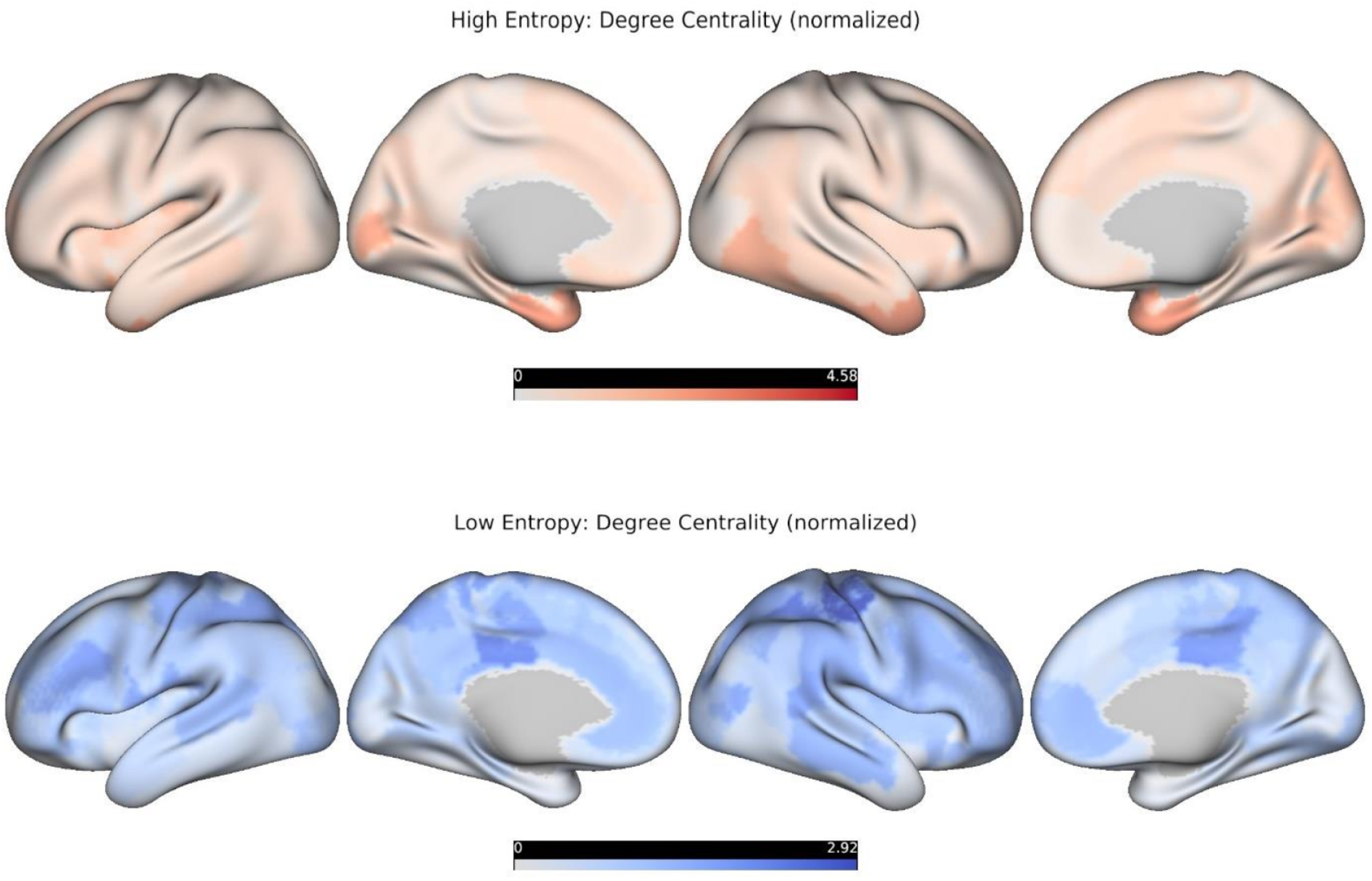
*<u>Top-row:</u>* Normalized degree-centrality of cortical nodes for High-entropy (red colormap), projected onto an inflated representation of the cortical surface. Darker colors denote higher normalized degree. Normalization was done via randomly shuffling edges (10000 permutations). Data are the same as in Figure 4. *<u>Bottom-row:</u>* Normalized degree-centrality of cortical nodes for Low-Entropy (blue colormap), projected onto an inflated representation of the cortical surface. Darker colors denote higher normalized degree. Normalization was done via randomly shuffling edges (10000 permutations). Data are the same as in Figure 4.

### 2.5 Topography of ESE mirrors macroscale patterns of cortical organization

We combined the normalized HEN/LEN DC estimates for each cortical node by subtracting them from each other (HEN - LEN) before rescaling them to the interval [0, 1] (Figure 7, top). The resulting node-entropy (NE) value captures a region’s trend towards being central in either HEN or LEN, at the behaviorally most informative density of these respective configurations (Section 2). NE values were then correlated with the corresponding values of a series of spatial maps denoting densities of different neurotransmitter receptors (from PET) and oscillatory power within predefined frequency bands (from MEG), see Hansen et al. (2022) for details (Figure 8). We restricted our analyses to maps for which the absolute Pearson correlation with NE was at least 0.2. For each map, significance was determined by comparing the empirical correlation value to a corresponding distribution derived from 10000 surrogate maps preserving the spatial autocorrelation of the initial map (Burt et al., 2020), and finally these p-values were controlled with FDR. We found that NE was significantly anticorrelated with MEG beta-power *(r=* -0.51,p = 0.001) and receptor density for the norepinephrine transporter (NET; *r= -0*.*4,p* = 0.012) (Figure 7). NE was also significantly correlated with receptor density for the serotonin transporter (5-HTT; *r* = 0.4, *p* = 0.012) (Figure 7). These results indicate that ESE at the node level tracks spatial gradients related to large-scale neuronal dynamics and neurotransmission. Importantly, they also offer valuable additional information to properly interpret the relationships between ESE and different cognitive domains in PSD we have described above.

**Figure 7.**
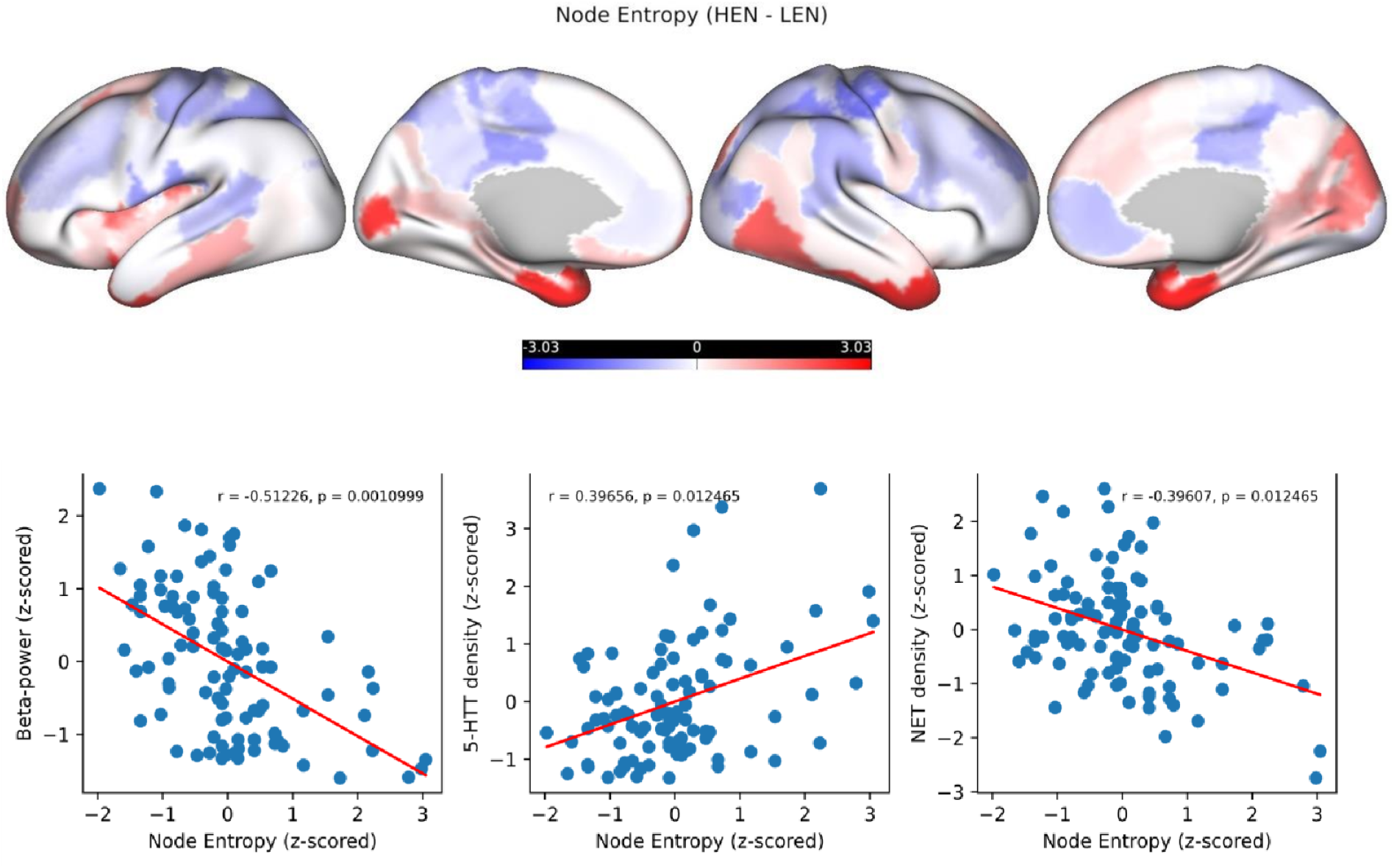
*<u>Top-row:</u>* Cortical Node-entropy, obtained by subtracting the normalized-degree centralities (High-entropy - Low-entropy), projected onto an inflated representation of the cortical surface and z-scored for visualization purposes. Darker red colors denote higher Node-entropy, darker blue colors lower Node-entropy. *<u>Bottom-row:</u>* Scatterplots depicting the relationship between Node-entropy (x-axis), and Beta-power (y-axis; left), 5-HTT density (y-axis; middle), and NET density (y-axis; right), for cortical nodes. Red lines denote least-square fits from linear regression. All variables were z-scored for visualization. P-values were derived with spatial-surrogate testing and then controlled with FDR (see Section 2.5). 5-HTT = Serotonin-transporter, NET= Norepinephrine-transporter.

**Figure 8.**
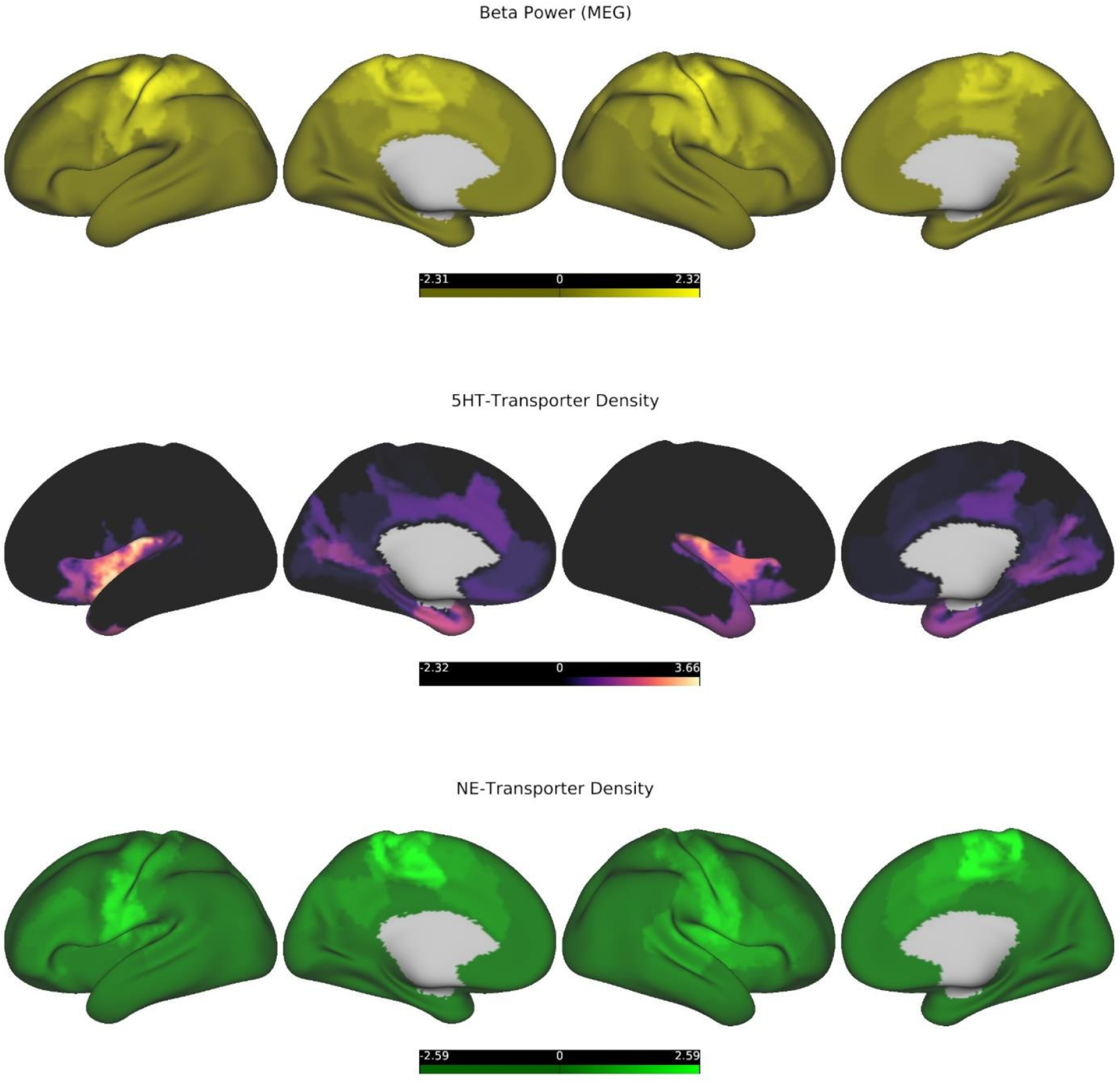
*<u>Top-row:</u>* MEG-beta power, projected onto an inflated representation of the cortical surface and z-scored for visualization purposes, lighter colors denote higher beta power. *<u>Middle-row:</u>* 5HTT-density, projected onto an inflated representation of the cortical surface and z-scored for visualization purposes, warmer colors denote higher density. *<u>Bottom-row:</u>* NET-density, projected onto an inflated representation of the cortical surface and z-scored for visualization purposes, lighter colors denote higher density. Data were taken from a public repository (see Section 4.1). 5-HTT = Serotonin-transporter, NET= Norepinephrine-transporter.

### 2.6 Additional analyses

#### 2.6.1 LEN maps integration

Our pattern of results is compatible with the hypothesis that LEN configurations preferentially encode behavioral information on tasks that require extensive information-integration (such as language- and knowledge-based tasks), while HEN tends to explain more variance on tasks geared more towards quick and reliable extraction of stimulus features (EF tasks measuring INH and WM). However, this a-priori grouping into cognitive domains (language vs. EF) is problematic and not clearly reflected in our findings, given that LEN explains the most variance in the EF ‘subdomain’ of Flexibility (Figures 1-2). To further test our hypothesis in a data-driven way, we conducted a principal component analysis (PCA) on the cognitive variables from the whole sample. The first principal component (PC), which explained approximately 52% of the variance, had positive loadings from all cognitive variables, indicating that it represented shared features across all tasks and their related domains (Figure 9, left). Interestingly, the average loading on the first PC was significantly lower for variables whose variance was significantly better explained by HEN (INH, WM), compared to variables that were more related to LEN (Flexibility, Reading, and Vocabulary comprehension), 95% BS-CI [-0.14; -0.02] (Figure 9, left). It seems that in terms of explanatory power, LEN configurations outperform HEN configurations, specifically on tasks that engage a wide range of cognitive domains, possibly accompanied by higher degrees of integrative and distributed processing on a neuronal level.

**Figure 9.**
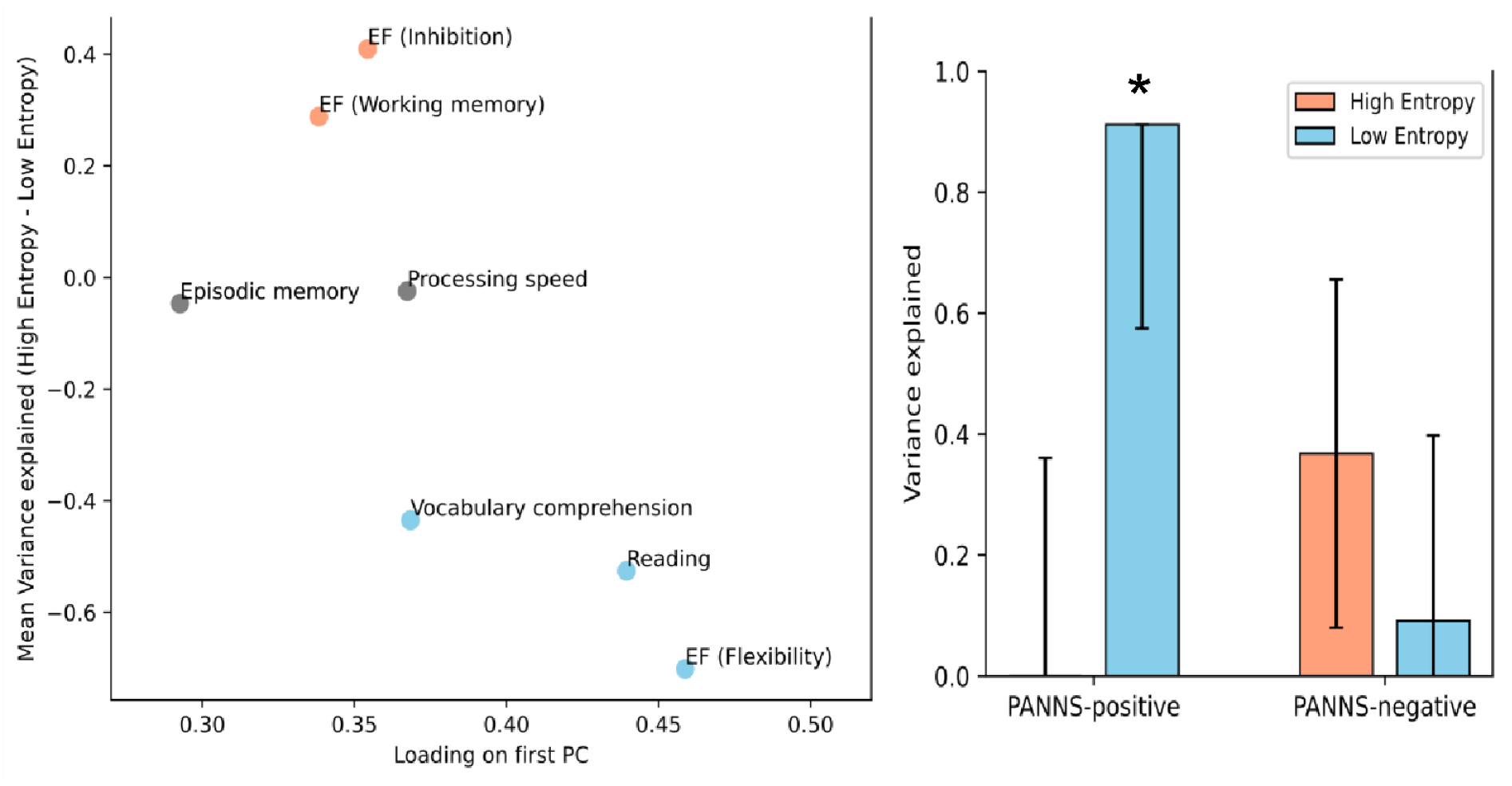
*<u>Left:</u>* Scatterplotvisualizing the relationship between integration (loading on first PC; x-axis) and difference in variance explained (High-entropy - Low-entropy; y-axis), stratified by cognitive measure. Measures are color coded according to direction and significance of the difference (blue: High-entropy< Low-entropy; red: High-entropy > Low-entropy; grey: ns.). *<u>Right:</u>* Low-entropy (blue) significantly explains variance in positive symptoms in patients. Error bars denote parametric SEs, significance is denoted by an asterisk. PC= Principal component, SE= Standard error.

#### 2.6.2 LEN relates to positive PSD pathology

While our main aims in this study were connected to cognitive variables, we also performed explorative analyses to see if and how HEN and LEN signatures would relate to the positive and negative symptom complexes that characterize PSD pathology. To do this we ran the univariate version of the statistical model (Section 4.5.1) with the patients scores on the Positive and Negative Syndrome Scale (PANSS) (Liechti, Capodilupo, Opler, Opler, & Yang, 2017) as dependent variables. The same covariates were used as in the analyses of the cognitive data. We found that LEN significantly encoded inter-patient variance on the positive scale (91%; *SE* = 34%; p-Wald = 0.0034), which was not the case for HEN (Figure 9, right). Additionally, there was some weaker evidence that HEN significantly explained variance on the negative symptom scale (37%; *SE* = 29%; p-Wald = 0.11) (Figure 9, right). Although these results should be interpreted with caution (Sabuncu et al., 2016), they show a correspondence between our suggested marker of neuronal integration (ESE) and core PSD symptoms like hallucinations and delusions, which fittingly have been hypothesized to stem from faulty integration of information between sensory- and higher order brain systems (Anticevic & Halassa, 2023; Friston et al., 2016).

## 3 Discussion

### 3.1 Summary

In this study, we show that the timescales of TVFC during rest encode significant information about cognitive task performance in a large sample of young adults in the early phases of psychosis. Our hypothesis was that diverging levels of ESE in patients would be differentially related to cognitive variables, depending on the level of integrative processing required for performing the related task. We find that brain configurations marked by low ESE (high integration; LEN) explain significantly more behavioral variance overall in patients, compared to constellations designated by high ESE (low integration; HEN). In line with our proposal, this result is driven by LEN encoding significantly more variance on tasks that engage a wider range of cognitive processes (Section 2.6.1). Fittingly, the most informative connections of the LEN are distributed across a range of cortical NWs encompassing unimodal-as well as higher-order cortical NWs (Section 2.3). In contrast, the most informative HEN connections are concentrated between SC and CC (Figure 3, bottom-left). This mostly reflects significantly higher relative explanatory power for HEN in tasks related to (empirically) more isolated EF subdomains (WM, INH). In sum, ESE is a useful marker for disentangling the relative contributions of different brain systems to specific aspects of cognitive performance in PSD.

In healthy controls, ESE decreases along a SC to cortical gradient and is lowest for nodes belonging to SM, DAT and CC, which replicates our previous findings (Hirsch & Wohlschlaeger, 2022). For cortical areas, NE closely covaries with oscillatory power in the beta frequency range during rest (ca. 12-30Hz), with lower NE corresponding to higher beta power (Figures 7-8). This points towards a possible neurobiological mechanism through which information could be integrated in the LEN, especially since NE also tracks the density of NET (Figures 7-8), with lower NE corresponding to higher density. Additionally, NE is significantly related to 5-HTT density, with higher NE corresponding to higher density (Figures 7-8). Overall, these results provide valuable insights into the oscillatory and neuromodulatory profiles of HEN and LEN. Given that the edge dynamics of PSD patients within these configurations are significantly related to their cognitive profiles, they also provide a mechanistic framework for possible interventions to improve cognition in PSDs. This is especially important, since our explorative analyses also show that LEN dynamics in patients are significantly related to positive symptoms like hallucinations and delusions (Figure 9, right). In the following sections we will discuss the implications of our findings in the context of the existing literature.

### 3.2 Relationships with previous work

The outcomes of the present investigation validate the substantial body of literature showing that resting-state dynamics are useful biomarkers for neuropathological conditions (L. G. Bauer et al., 2022; Kaiser et al., 2016; Rashid et al., 2016; Ries et al., 2019; Sakoglu et al., 2010; Salman, Vergara, Damaraju, & Calhoun, 2019; Zöller et al., 2019), and that ESE significantly relates to behavior and cognition (Jia & Gu, 2019a; Jia et al., 2017; S.S. Menon & Krishnamurthy, 2019). However, our detailed mapping of HEN/LEN configurations and their multimodal profiles to specific aspects of cognition and positive pathology in PSD provides new theoretical insights and has possible clinical utility.

#### 3.2.1 LEN

NWs significantly related to cognition in LEN include DMN, SAL and CC, all of whom are part of the triple-network model of general psychopathology (V. Menon, 2011). The model postulates that cognitive deficits in SCZ and psychopathology in general arise from dysfunctional interactions between these higher order NWs (V. Menon & Uddin, 2010; Palaniyappan & Liddle, 2012). Indeed, their dysfunction is predictive of cognitive deficits across modalities and diagnostic criteria (Sheffield et al., 2017; Sui et al., 2018), which also holds true in the present investigation. Since the LEN/HEN templates were derived from group-average ESE values across healthy individuals (Section 2), our results partially reflect the spatial gradient of ESE (Figures 4-6), with nodes belonging to these higher order NWs amid the most highly connected in the LEN. However, the behavioral significance of the patients LEN dynamics was entirely absent when edges were selected randomly (Section 2.1). This suggests that the TVFC timescales within and between those NWs were indeed amongst the most informative about specific aspects of cognition. The lack of significance for any NW or NW-interaction after controlling for degree and strength (Figure 3, bottom-right) indicates that the behavioral relevance of LEN is not so much concentrated but rather distributed across its constituent nodes and associated NWs. This conceptually aligns with our finding that LEN dynamics preferentially encode performance in cognitive tasks requiring higher degrees of integration.

The fact that the topography of NE was strongly anticorrelated with MEG beta-power (Figure 7) implies coordinated activity between nodes within the LEN, underscoring that this configuration is not merely incidental. Beta power has been related to ongoing effortful cognition (Schmidt et al., 2019) and motor preparation/execution (Baker, 2007; Pfurtscheller & Berghold, 1989; Tewarie et al., 2018). Interestingly, this rhythm seems to be important for integrating bottom-up and top-down signals (Tan, Wade, & Brown, 2016), and tracks SCZ pathology (Donati et al., 2021; Gascoyne et al., 2021; Pittman-Polletta, Kocsis, Vijayan, Whittington, & Kopell, 2015). LEN dynamics were also significantly related to positive symptom severity in the present study (Figure 9, right), providing further evidence for a possible link between ESE and large-scale neuronal dynamics. The NE connection to oscillatory behavior should be interpreted together with the corresponding spatial correlations to NET and 5-HTT densities (Figure 7). Recent evidence shows that these distributions significantly predict the topography of MEG-derived beta-power (Hansen et al., 2022), and monoaminergic dysfunction is central to many explanatory accounts of PSDs (Davis, Kahn, Ko, & Davidson, 1991; Eggers, 2013). The noradrenergic system has been implicated in the cognitive deficits in SCZ patients (Maki-Marttunen, Andreassen, & Espeseth, 2020), and has been hypothesized to drive integration between distributed brain NWs through neural gain (Shine, 2019; Totah, Neves, Panzeri, Logothetis, & Eschenko, 2018). Since NE was significantly anticorrelated with NET density, these accounts align with our notion that ESE inversely tracks integration, behaviorally reflected in the (relatively) superior performance of patients’ LEN patterns to encode variance in psychometrically more integrated tasks, which evidence suggest require higher degrees of distributed processing (Colom, Jung, & Haier, 2006; Dajani & Uddin, 2015; Niendam et al., 2012).

A corollary of our results is that psychoactive interventions that target positive symptoms in PSDs should also significantly influence cognitive performance, given that LEN dynamics were significantly related to both aspects of the pathology. There is indeed evidence that some antipsychotics have small positive effects on cognition (Baldez et al., 2021; Davidson et al., 2009), with negative effects also being reported (Sakurai et al., 2013). Of note, a recent network meta-analysis showed that the antipsychotics haloperidol and clozapine, which are known for their antagonistic effects on noradrenergic transmission, had the most detrimental effects on global cognition (Baldez et al., 2021). This is compatible with our present results that indicate a noradrenergic involvement in the LEN dynamics which significantly encode cognitive-task variance in PSD patients. Interestingly, NET can also modulate dopaminergic (DA) signaling, especially in CC related areas (Gresch, Sved, Zigmond, & Finlay, 1995; Maki Märttunen et al., 2020; Moron, Brockington, Wise, Rocha, & Hope, 2002), and DA dysfunction has been the central element in many theories of PSDs (Howes & Kapur, 2009).

#### 3.2.2 HEN

Areas significantly related to behavior in the HEN pertained to interactions within SC, and SC-interactions with VIS and higher-order NWs (Figure 3, top/bottom-left). These associations were driven by HEN explanatory power in specific tasks (List sorting and Flanker) related to EF subdomains (Working memory [WM] and Inhibition [INH]). These tasks require quick and precise encoding oflow-levels stimulus features to perform well (Tulsky et al., 2013; Zelazo et al., 2013). Our results suggest that connections with high ESE (low integration) best encoded this ability during rest in PSD patients, which is compatible with our hypothesis. This is in line with evidence that SC and visual areas have shorter INTs, compared to cortical higher-order areas (Muller et al., 2020; Raut et al., 2020), which is also true for the timescales of TVFC (Hirsch & Wohlschlaeger, 2022). Interactions within SC are proposed to act as shortcuts for rapid sensory processing (Mcfadyen, Dolan, & Garrido, 2020), and SC-cortical interactions have been consistently associated cognitive symptoms of PSDs (Anticevic & Halassa, 2023; Peters, Dunlop, & Downar, 2016; Ramsay, 2019), possibly by influencing cortico-cortical connectivity (Hirsch & Wohlschlaeger, 2023). The behaviorally most informative HEN interactions were concentrated between SC and CC (Figure 3, bottom-left), contrasting the more distributed nature of relevant LEN edges. In general, ESE was able to dissociate different aspects of EF (WM/INH vs. Flexibility) in terms of their neurophysiological correlates, resembling the *stability* vs. *flexibility* dichotomy of cognitive control (Fuster, 2015; Sakai, 2008).

The observed significant correlation between 5-HTT density and NE for cortical nodes indicates an involvement of the serotonergic system in HEN dynamics (Figure 7), especially since FC changes after 5-HTT blockage have been reported for HEN regions including the thalamus, amygdala, and VIS (Boucherie et al., 2023). Moreover, serotonergic signaling under normal conditions has been related to (SC-driven) feedforward cortical processing (Shine et al., 2022), which is associated with shorter timescales (Bastos et al., 2012). Of note, performance in WM and selective attention (akin to INH) was improved for PSD patients after administration of the AP olanzapine, relative to other atypical APs, typical APs, and placebo (Baldez et al., 2021; Neil D Woodward, Purdon, Meltzer, & Zald, 2005). These improvements were partially attributed to olanzapine’s increased affinity for some serotonergic receptors (Baldez et al., 2021; Bymaster et al., 2001; Neil D Woodward et al., 2005), aligning with evidence showing serotonergic effects on WM (Williams, Rao, & Goldman-Rakic, 2002) and INH (Pattij & Schoffelmeer, 2015). Collectively, these findings suggest a neurobiological basis for our observed relationship between HEN timescales and specific aspects of cognition in PSD.

### 3.3 Limitations

While our hypothesis was based on the notion from INTs that more self-similarity indicates a greater potential for integration (Hasson et al., 2015; J. D. Murray et al., 2014), ESE is only indirectly related to the BOLD signal via TVFC. However, our findings in PSD patients indeed suggest that TVFC configurations marked by more regular fluctuations (low ESE) explain variance better on tasks that require more integrated processing (Figure 9). TVFC fluctuations have been interpreted as shifting brain-states (Allen et al., 2014; Leonardi & Van De Ville, 2015), reflecting underlying electrophysiological phenomena (Tagliazucchi, Von Wegner, Morzelewski, Brodbeck, & Laufs, 2012; G. J. Thompson, 2018) and neuromodulatory processes (Shafiei et al., 2019; Shine, 2019), which is compatible with our findings. A criticism of our methodology could be that ESE might not be sensitive to active communication between two given regions. It is certainly possible for an edge to have low ESE (high integration) but for the two corresponding nodes to have low or negative FC. However, we do not think that such connections should be excluded or that their existence invalidates our interpretation of ESE. On the contrary, evidence shows that weak connections are especially informative about cognition (Santarnecchi, Galli, Polizzotto, Rossi, & Rossi, 2014), and topology in PSD (Bassett, Nelson, Mueller, Camchong, & Lim, 2012; Mastrandrea et al., 2021).

Another possible issue is the fact that SampEn (by definition) is influenced by basic signal properties like temporal signal-to noise ratio (Keilholz et al., 2020), which is lower for BOLD signals from SC and temporal regions. Although we have shown in the past that the implications for ESE are small (Hirsch & Wohlschlaeger, 2022), these influences must be kept in mind when interpreting spatial patterns of ESE. Nevertheless, the observed relationships between ESE at different levels and behavior, oscillatory dynamics and neurotransmitter densities suggest relevant information can be extracted from the timescales of TVFC. Finally, in the past we and others have equated high (single-scale) SampEn with high complexity (Hirsch & Wohlschlaeger, 2022; Jia & Gu, 2019a, 2019b; Jia et al., 2017), but others have argued that such an interpretation requires a multi-scale entropy analysis (Costa, Goldberger, & Peng, 2002; A. C. Yang et al., 2015). While we have avoided the notion of complexity in the present study, it should be noted that contrary to BOLD SampEn, ESE at our scale of interest captured most of the behaviorally relevant information in healthy subjects (S. S. Menon & Krishnamurthy, 2019).

### 3.4 Clinical implications

Our finding’s main (potential) clinical utility lies in the association of distinct aspects of cognition in PSD with the topography of neurotransmitter- and oscillatory systems, via ESE. Although (small) positive effects of APs on cognition have consistently been reported (Baldez et al., 2021; Keefe et al., 2007), our findings suggest that pharmacological interventions specifically aimed at noradrenergic and/or serotonergic systems might proof beneficial in terms of improving specific aspects of cognition in PSD and related disorders. While some evidence exists in that regard (Mancini et al., 2021; Silver et al., 2015), no clinical relevance of antidepressants in general was reported in a recent metanalysis of chronic SCZ patients (Vernon et al., 2014). However, the included studies were small, and cognitive outcomes were grouped within a-priori cognitive domains (EF, language, etc.) (Vernon et al., 2014). Our results suggest that such grouping could obscure possible effects. In addition, our findings pertain to young patients in the early phases of PSD, not chronic SCZ. Finally, the implication of the beta-rhythm in the LEN makes it a potential target of brain-stimulation techniques, which is technically feasible with non-invasive methods (Hannah, Muralidharan, & Aron, 2022).

## 4 Matetials and Methods

### 4.1 Sample characteristics and image preprocessing

The initial sample consisted of the 169 subjects for which minimally preprocessed structural data was available at the time of download as part of the Human Connectome Project Early Psychosis Release 1.1 (HCP-EP) (https://www.humanconnectome.org/study/human-connectome-project-for-early-psychosis/document/hcp-ep). For these subjects the (volumetric) minimal preprocessing pipeline of the HCP was conducted, see (Glasser et al., 2013; Smith et al., 2013) for details. Briefly, one resting-state fMRI run lasted 5min and 47s, 2 mm isotropic resolution, multiband acceleration factor 8, TR= 0.8s, TE= 0.037s, phase-encoding direction posterior-to-anterior (PA). Additional runs were available in the anterior-to-posterior (AP) phase-encoding direction, but we only used one run with PA per subject to ensure better signal accuracy in frontal regions. Preprocessing delivered unsatisfying results for five subjects due to issues with the field-maps, which were subsequently excluded from further analysis. Out of the remaining 164 subjects, 150 subjects had sufficient behavioral data available (patients: *n* = 97; controls: *n* = 53), which were then included in the final sample. Functional data were then denoised with aCompCor (Behzadi, Restom, Liau, & Liu, 2007), consisting in regressing out the timeseries of the five main axes of variance (principal components) from white-matter and cerebrospinal signals (respectively) from the functional images (Muschelli et al., 2014). Additionally, the six movement parameters and their derivatives were regressed out, and the images were downsampled to 116 cortical and subcortical (SC) regions (Tian, Margulies, Breakspear, & Zalesky, 2020), with a template from (https://github.com/yetianmed/subcortex/blob/master/Group-Parcellation/3T/Cortex-Subcortex/MN!volumetric/Schaefer2018_l00Parcels_7Networks_order_Tian_Subcortex_Sl_MNil_52NLin6Asym_2mm.nii.gz). Preprocessed PET *(n =* 19) and MEG *(n =* 6) spatial maps were obtained from a public repository (https://github.com/netneurolab/hansen_receptors) at the 100 parcel resolution of the Schaefer atlas (Schaefer et al., 2018).

### 4.2 SWA and ESE calculation

Prior to SWA, data were bandpass filtered between 0.017 Hz and 0.1 Hz (Leonardi & Van De Ville, 2015), and the mean signal across all regions was regressed from the data (a version of global-signal regression [GSR]). GSR has been shown to be beneficial for alleviating the influence of global artifacts in fMRI data (Burgess et al., 2016), strengthen brain-behavior relationships on task measures (Li et al., 2019), and increases sensitivity to FC differences between controls and clinical populations (Parkes, Fulcher, Yücel, & Fornito, 2018). The first/last 10 frames were removed to account for any boundary effects. We used a rectangular window with a width corresponding to 60s, which was then slid in steps of one TR across the timeseries. Within each window we estimated the Pearson correlation between all regions, which was then Fisher-transformed prior to further analysis. Then SampEn was then calculated for each correlational timeseries. To compute the SampEn for a given signal *x* = [ *x*_*1*_, *x*_*2*_,…, *x*_*n*_] with length *N*, an embedding vector with *m* running data points is derived from x: *V*_*i*_ = [*x*_*i*_, *x*_*i+1*_, …, *x*_*i+m-1*_], with *m* corresponding to the embedding dimension. For each *i* (1*≤i ≤N*−*m*) define

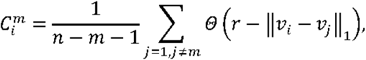

where *r* = εσx corresponds to a tolerance value, ε to a scaling parameter, and σx to the standard deviation of *x*. Θ(·) is the Heaviside function

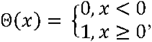

and ‖ ·‖ _1_ is the Chebyshev distance

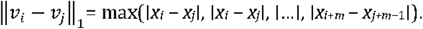

Then, for each *i* (1*≤i ≤N*−*m*) define

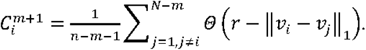

Averaging over all *m* embedding vectors gives

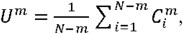

and

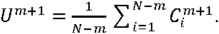

SampEn is defined as

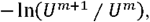

resulting in a nonnegative number, with higher values indicative of less regularity in the signal (Richman & Moorman, 2000). To ensure comparability of our results with past investigations, we used the standard parameter values of *m =* 2 and *ε* = 0.20 (Hirsch & Wohlschlaeger, 2022; Jia & Gu, 2019b). For BOLD signals of at least 97 timepoints evidence suggests that results from SampEn analyses are robust to parameter changes (Albert C. Yang, Tsai, Lin, & Peng, 2018), and similar results were obtained for ESE (Jia et al., 2017).

### 4.3 Construction of HEN/LEN templates and similarity matrices

After ESE values were obtained for all subjects, HEN and LEN templates were constructed by proportional thresholding of the ESE matrix averaged across healthy individuals. For each cutoff only a certain proportion of the highest (HEN) or lowest (LEN) edges was kept. The resulting 32 templates (16 HEN and 16 LEN) were then used as binary masks to extract the corresponding ESE values from the patients, which were then correlated across patients to obtain the similarity matrices. We then ran the statistical model (Section 4.5.1) for each cutoff with the corresponding similarity matrices as inputs. Each cutoff was ranked according to explanatory power (mean variance explained), significance (*p*-Perm and *p*-Wald), and accuracy (absolute difference between *p*-Perm and *p*-Wald, see Ge et al. (2016) for the rationale). Subsequently, the average rank across all criteria was calculated and the cutoff with the highest rank was chosen for all downstream analyses. For HEN the optimal cutoff was at ∼6% density and for LEN at ∼14% density. Importantly, the complexity of the models for HEN and LEN is equivalent, since the final model inputs (the similarity matrices *RHEN* and *RLEN)* have equal dimensions, see Liegeois et al. (2019) for a discussion.

### 4.4 Behavioi·al variables

#### 4.4.1 Cognitive variables

The seven selected behavioral variables constitute the cognitive module of the NIH Toolbox for the Assessment of Neurological and Behavioral Function, which measures the cognitive domains of EF, episodic memory, language, processing speed, working memory, and attention (Weintraub et al., 2013). Under EF we grouped the subdomains of Flexibility (Dimensional Change Card Sort), Inhibition (flanker task), and Working memory (list sorting working memory test) (Tulsky et al., 2013; Zelazo et al., 2013). Language functions were denoted by Reading (Oral Reading Recognition Test) and Vocabulary comprehension (Picture Vocabulary Test) (Gershon et al., 2013), and Episodic memory was assessed with the Picture Sequence Memory Test (P. J. Bauer et al., 2013). Finally, Processing speed was assessed with the Pattern Comparison Processing Speed Test (Carlozzi, Tulsky, Kail, & Beaumont, 2013). For all tests the age-corrected scaled scores were utilized (Weintraub et al., 2013). One subject had a missing score for Episodic memory, which was set to the median value across subjects. Prior to being entered into the model, the variables were quantile normalized to a Gaussian distribution to fit model assumptions (Liegeois et al., 2019).

#### 4.4.2 Covariates

We included age, sex, phenotype description (non-affective vs. affective psychosis), current medication (Chlorpromazine equivalents), as well as mean framewise-displacement (Power, Barnes, Snyder, Schlaggar, & Petersen, 2012) as covariates in the model. One subject had a missing current-medication value, which was set to zero, given that the subjects lifetime exposure to antipsychotic medication was denoted by zero.

### 4,5 Variance component model

#### 4.5.1 Multivariate model

The multidimensional variance component model of Ge et al. (2016) takes the following form:

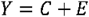

where *Y, C*, and *E* are 97 x 7 matrices, with *Y* representing the (quantile normalized) cognitive variables for all *N* subjects (Section 4.4.1). *Vec*(*C*) ∼*N*(0,∑_*c*⊗_*R*), and *Vec*(*E*)∼*N*(0,∑_*e*⊗_*I*), where *Vec*(.) is the vectorization operator, ⊗ the Kronecker matrix product, *R* the similarity matrix (i.e., either *RHEN* or *RLEN)* and *I* the identity matrix. The 97 x 97 matrices ∑_*c*_ and ∑_*e*_ are to be estimated from *R* and *Y*, which can be done with a moment-matching method (Ge et al., 2016):

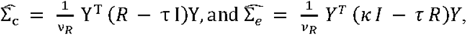

where 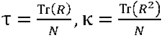 and *v*_*R*_ = *N*(*κ* - τ^2^). The overall behavioral variance across all measures *M* (explained by either HEN or LEN), is then:

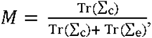

with **Tr(.)** being the trace operator. The explained variance for a single cognitive variable *M*_*i*_; is computed as:

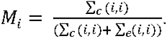

This results in one value between zero and one, across all measures, and for each behavioral measure. Since we account for covariates, the model becomes:

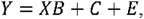

where *X* is the 97 x 5 matrix of covariates (Section 4.4.2), and *B* a 5 x 7 matrix of fixed effects (Ge et al., 2016). To remove the covariate matrix from the model, the data is projected onto a 97 − 5-dimensional subspace resulting in the transformed model:

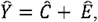

which is equivalent to the original model, see Ge et al. (2015) for details. Significance for the average variance across all cognitive variables was measured with a *P* value derived from a Wald test (*p*-Wald), and complementarily by permuting the rows and columns of *R* (*p*-Perm) (Ge et al., 2016). The results described in Section 2.1 were calculated by running the model separately for *RHEN* and *RLEN*, as well as for their alternatively derived (random) versions (see Sections 2.1 and 4.3).

#### 4.5.2 Univariate model

To obtain the results described in Section 2.3 we used a univariate version of the multivariate model (Liegeois et al., 2019; Sabuncu et al., 2016):

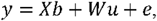

where *y* corresponds to a vector whose *N* entries contain the values of a given cognitive variable for all subjects, and *b* to a vector of fixed effects. *W* is a column-standardized *N* x *P* matrix, with *P* corresponding to the total number of edges in the HEN/LEN template (see Section 4.3), *u* ∼ *N*(0, σ_*c*_ / *P)* is a vector of random effects, and *e* ∼ *N(0, σ*_*e)*_ the residual. *W* contains the subjects’ standardized SampEn estimates for a given template, and assuming that the elements of *u* are independent, this model can be transformed to the model from Section 4.5.1:*Cov*(*y*) = *σ*_*c*_ · *R* + *σ*_*e*_ · *I*, with *R* = *W·W*^*T*^*/P* The squared entries of the best linear unbiased predictor of *u*(*û*) (J. Yang, Lee, Goddard, & Visscher, 2011) are a scaled estimate of the variance explained by the corresponding edge across all individuals. We computed *û*^2^ for each cognitive variable and weighted it by the loading of that variable on the first principal component derived from the whole set of behavioral variables (Liegeois et al., 2019), see Ge et al. (2016) for the underlying rationale.

### 4.6 Statistical inference

The significance of the results reported in Section 2.2 was assessed by estimating CIs after resampling, since the model output consist of just one value between zero and one for every dependent variable, as well as the average variance explained. We used two different resampling methods to ensure robustness of the ensuing CIs: BS-Cls were calculated based on 1000 bootstrap samples and their width was Bonferroni adjusted for multiple comparisons (Manly, 2018). As a complementary approach, we employed a block-version of jackknife-resampling in combination with random subsampling: Briefly, the JK-CIs were computed based on 1000 subsets of size *N*− *d* of the original data, randomly sampled with replacement (Shao&Tu, 1995a). If 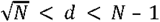, this yields a consistent jackknife variance estimator for most underlying statistics, so we used *d =* 10 (Shao & Tu, 1995b).

The significance of the results reported in Section 2.3 was assessed with two different null-models that were derived from the results of the univariate analysis (see Section 4.5.1). For both HEN and LEN (respectively) these results can be represented by a graph with 116 nodes whose edges are weighted according to the average behavioral variance explained across variables. The binarized versions of these graphs are equivalent to the corresponding HEN/LEN template. For the first null-model we just randomly shuffled the edges (10000 iterations) before calculating the average variance explained at the system-level (see Section 2.3). However, we also wanted to know if there were certain NWs/NW-interactions that explained significantly more (average) behavioral variance than would be expected, based on the relative centrality and explanatory power of their constituent nodes in the underlying templates. To achieve this we created degree- and strength matched random NWs (10000 iterations) (Rubinov & Sporns, 2011), before calculating the average variance explained at the system-level.

### 4.7 Softwue and code used in the analysis

aCompCor denoising was done with DPABI (Yan, Wang, Zuo, & Zang, 2016), which was developed in MATLAB (The MathWorks Inc., Natick, MA, US). Further preprocessing was done in Python with the help of the Nilearn toolbox (https://zenodo.org/records/10579570). SWA was done in Python with TENETO (https://zenodo.org/records/3626827) (W. H. Thompson, Brantefors, & Fransson, 2017). SampEn calculation was done in Python with EntropyHub (Flood & Grimm, 2021). Further statistical analyses were done in MATLAB with the help of the following toolboxes: Brain Connectivity Toolbox (BCT) (https://sites.google.com/site/bctnet/home). statistics-resampling package (https://doi.org/10.5281/zenodo.3992392), and the BrainSpace toolbox (https://brainspace.readthedocs.io/en/latest/index.html) (Vos de Wael et al, 2020). Figures were in part created with Matplotlib (Hunter, 2007), MNE (https://doi.org/10.5281/zenodo>.592483) (Gramfort et al., 2013), NiBabel (https://zenodo.org/records/10363247), and Connectome Workbench (Marcus et al., 2011). Custom code and further materials associated with the study can be found here *(-link-to-be-added-)*.

## Data Availability

Research using Human Connectome Project for Early Psychosis (HCP-EP) data reported in this publication was supported by the National Institute of Mental Health of the National Institutes of Health under Award Number U01MH109977. The HCP-EP 1.1 Release data used in this report came from DOI: 10.15154/1522899. Further materials associated with the study will be made available upon publication.

https://www.humanconnectome.org/study/human-connectome-project-for-early-psychosis/document/hcp-ep

## 4.8 Aclmowledgements

Research using Human Connectome Project for Early Psychosis (HCP-EP) data reported in this publication was supported by the National Institute of Mental Health of the National Institutes of Health under Award Number U01MH109977. The HCP-EP 1.1 Release data used in this report came from DOI: 10.15154/1522899.

